# Quality of Refractive Error Care in Cambodia: An Unannounced Standardised Patient Study

**DOI:** 10.1101/2023.06.11.23291172

**Authors:** Anthea Burnett, Ngy Meng, Do Seiha, Neath Kong, Seila Chea, Malis Dean, Piseth Horm, Kim San Meas, Beatrice Varga, Suit May Ho, Myra McGuinness, Ling Lee

## Abstract

**Purpose:** Quality-of-care in refractive error services is essential, as it directly affects vision outcomes, wellbeing, educational attainment, and workforce participation. In Cambodia, uncorrected refractive error is a leading cause of mild and moderate vision impairment in adults. We evaluated the quality of refractive error care in Cambodia by estimating the proportion of prescribed and dispensed spectacles appropriate for people’s refractive error needs and factors associated with spectacle quality.

**Methods:** A cross-sectional protocol was employed with 18 Khmer-speaking adult participants observing testing procedures in 156 optical services across six provinces in 2022. A total of 496 dispensed spectacles were assessed against spectacle quality indicators.

**Results:** The analysis revealed that 35.1% of dispensed spectacles were of optimal quality. The most common error observed in sub-optimal spectacles was the presence of horizontal prism outside of tolerance limits. The study also found that 44.0% of emmetrope visits involved unnecessary prescription spectacle recommendations, and 18.3% of written prescriptions did not correspond with dispensed spectacles. Sex differences were observed, with men predominantly providing refractive error care and women more likely to be unnecessarily recommended prescription spectacles.

**Conclusion:** The findings highlight the importance of prioritizing quality-of-care in refractive error services. A key recommendation is to consider regulatory mechanisms to ensure optical services employ appropriately qualified staff. Additionally, efforts should be made to eliminate unnecessary prescriptions –– especially for emmetropes and females –– standardize written prescriptions, ensure consistent pupil distance measurements, reduce reliance on autorefraction, and address the gender imbalance in the refractive error workforce.

## Introduction

Quality-of-care is a fundamental component within the context of universal health coverage (UHC), as it ensures that individuals receive effective, safe, and timely health care tailored to their needs, without experiencing financial hardship.[1] In terms of refractive error care, the emphasis on quality-of-care is significant, as it directly impacts visual outcomes, as well as wellbeing, educational attainment, and workforce participation.[2] High-quality refractive error care involves accurate diagnosis, appropriate prescription, and precise spectacle dispensing, all of which contribute to optimal visual acuity (VA) and comfort. By prioritising quality-of-care in refractive error services, healthcare providers can fully address the needs of patients with varying degrees of visual impairment.

In Cambodia, uncorrected refractive error causes 61% of mild vision impairment (presenting VA <6/12 – 6/18), and 17.5% of moderate vision impairment (presenting VA <6/18 – 6/60) in adults over 50 years.[3] In adults over 19 years, the estimated prevalence of myopia, hyperopia, and astigmatism is 49.1%, 4.45%, and 9.1% respectively,[4] and 21.5% of adults over 50 years live with near vision impairment.[5] In Takeo province, it was reported that more men than women (35% vs 10%) wear glasses, with the majority (68%) of glasses purchased from the market, 26% from an optical shop, 16% given from a relative, and 10% from Takeo Eye Hospital.[6]

Effective refractive error care (eREC) is one of two global targets endorsed by Member States at the 74th World Health Assembly to measure progress towards achieving UHC.[7] This target, which entails a 40-percentage point increase in eREC, is designed to monitor both the effective coverage and quality of refractive error care. Despite the absence of eREC estimates in Cambodia, it is anticipated that, significant enhancements in the quantity and quality of refractive services will be necessary to meet this global target. Therefore, it is crucial to identify and address the specific challenges and gaps in refractive error care within Cambodia to ensure progress towards UHC and improved eye health for all.

Unannounced Standardised Patients (USPs), covertly impersonate patients, and are considered the gold standard for assessing quality in clinical practice.[8] Their usage ensures an unbiased assessment of clinical techniques and services, as healthcare providers don’t alter their behaviors, not knowing they’re under observation. USPs have been instrumental in evaluating family planning, pharmaceutical dispensing, and clinical prescribing patterns in lower to middle income settings,[9] and even in assessing refractive error outcomes.[10-12]

Refractive error care encompasses identification of patient needs, a precise refraction, and dispensing spectacles that meet the patient’s refraction. A Quality of Refractive Error Care (Q.REC) study leverages USPs to generate evidence on the standard of delivered refractive services, informing potential alterations to practices and policies.

The aim of this study was to evaluate the quality of refractive error care in Cambodia by using USPs to estimate the proportion of prescribed and dispensed spectacles appropriate for people’s refractive error needs. Secondly, this study also aimed to assess the optical service and USP characteristics associated with spectacle quality and explore unnecessary prescribing practices.

## Material and Methods

This study employs a previously published cross-sectional protocol[13] to recruit 18 fluent Khmer-speaking adult USPs with either:

- Myopia: Spherical equivalent < -0.50 DS in at least one eye
- Hyperopia: Spherical equivalent > +0.50 DS in at least one eye
- Astigmatism: > 0.50 DC in at least one eye
- Emmetropia: Spherical equivalent ≥ -0.50 DS and ≤ +0.50 DS in both eyes
- Presbyopia: ≥ 1.00 DS above the best optical distance correction

USPs were excluded if they did not have good ocular health, had a history of refractive surgery, had manifest or intermittent strabismus or amblyopia, or any health conditions that could impact refractive error consistency.

The sampling frame for eligible optical services was compiled by collaborating with relevant Ministries of Health, Optical Councils, Optometry Associations, as well as cross-referencing with Google Maps. Optical services were deemed eligible if they provided refraction and dispensing services, were not previously known to USPs and did not have staff who worked across multiple services already selected.

A total of 202 optical services in Phnom Penh, Kandal, Siem Reap, Battambong, Kampong Cham and Takeo provinces were selected. However, as some services had relocated or closed due to COVID-19, only156 stores were visited. Service owners received written information about the study along with opportunities to opt-out verbally or in writing.

USPs were trained to visit stores in a standardised manner,[13] and received baseline refractions from three qualified local optometrists. USPs visited optical services, observed testing procedures and obtained one pair of single-vision spectacles, distance or near depending on the clinician’s recommendation. Research optometrists then assessed VA, comfort and lens prescriptions. Non presbyopic USPs with emmetropia did not purchase spectacles unless there was a risk of them being detected. This study follows the Checklist for Reporting Research Using Simulated Patient Methodology (CRiSP).[14]

### Quality outcomes

Spectacle quality was categorised as optimal or sub-optimal (Supplementary 1). Optimal quality single-vision spectacles met all quality components in both lenses, with the baseline distance sphere power plus any near addition for single vision near spectacles. Cylindrical axis quality was only assessed for USPs with cylinder power detected at baseline. Induced horizontal prism was derived using Prentice’s rule, from the lens power at horizontal meridian and the amount of lens decentration from the averaged baseline pupillary distance. Vertical prism was derived from focimetry.

The written prescription obtained from each visit was classified as optimal or sub-optimal relative to the baseline refraction using the tolerance limits for spherical and cylinder power and cylinder axis components. Dispensed spectacles matched the written prescription if spherical and cylindrical power were within 0.25 D, and cylinder axis tolerance limits were met. Unnecessary prescribing occurred when USPs with emmetropia and no presbyopia were advised to purchase spectacles.

VA with dispensed spectacles was considered good if it was within 1.5 lines of monocular and binocular baseline best-corrected VA. USPs reported any discomfort or eye strain while wearing the spectacles.

### Data analysis

Analyses were conducted using Stata/BE v16.0 (StataCorp LLC, College Station, TX). All dispensed spectacles were included in primary analysis. Secondary analyses examined associations between USP/optical service characteristics and spectacle quality.

Descriptive statistics were reported for baseline USP and optical service characteristics. Proportions were calculated for each outcome of interest (spectacle and prescription quality, comfort, spectacle-corrected VA, and unnecessary prescribing). Logit-transformed 95% confidence intervals (CI) were estimated using robust standard errors to account for intra-service correlation.

Univariable logistic regression was used to examine associations between USPs, optical service characteristics and spectacle quality. Multivariable models evaluated associations between service procedures (focimetry, autorefraction, distance and near subjective refraction) and spectacle quality, adjusting for USP baseline refraction type. Covariates were selected as potential confounders based on subject matter knowledge and the number of observations in each category. To account for within-optical service correlation, robust standard errors were used.

To account for the unequal numbers of spectacles dispensed in each province, weighting was applied so that pooled estimates could be generalized. Weights were assigned based on 2019 census population sizes, with Battambong, Kampong Cham, Ta Keo, Phnom Penh, and Siem Reap contributing 16.5%, 14.9%, 15.0%, 36.8%, and 16.7%, respectively. For each analysis set, each pair of spectacles within a province was weighted equally. As the primary aim of the study was to estimate the proportion of optimal-quality spectacles, comparative statistics were considered exploratory and no adjustment for multiple testing was applied.

## Results

This study analysed 16 USPs, consisting of 9 women and 7 men with ages ranging from 20 to 52 years. At baseline, all USPs had a best-corrected VA of logMAR 0.04 (∼6/6 part) or better for distance and 0.2 (∼6/9.5, N4) or better for near vision in each eye. The baseline spherical equivalent refraction for distance ranged from -5.31D to +1.04D, with a maximum of 1.25D cylinder power. The average interocular difference in spherical equivalent was 0.37D (SD 0.78, range 0.00 to 3.29D).

Each USP visited between 23 and 45 optical services, with each service being visited by up to five USPs (60% of services were visited by five USPs, 28% of services were visited by four USPs, 11% were visited by three USPs and the remaining services were visited by either one or two USPs). A total of 663 visits were made to 156 optical services, with 544 spectacles dispensed. After excluding ineligible visits (seven where the USP reported detection; eight visits to duplicate services; and 33 visits where a USP was later diagnosed with high Hba1c levels, which could have affected refractive error during data collection), 496 spectacles from 615 visits were included. Figure 1 depicts a flowchart of optical services, USPs, and spectacles included in the analysis.

**Figure 1:**
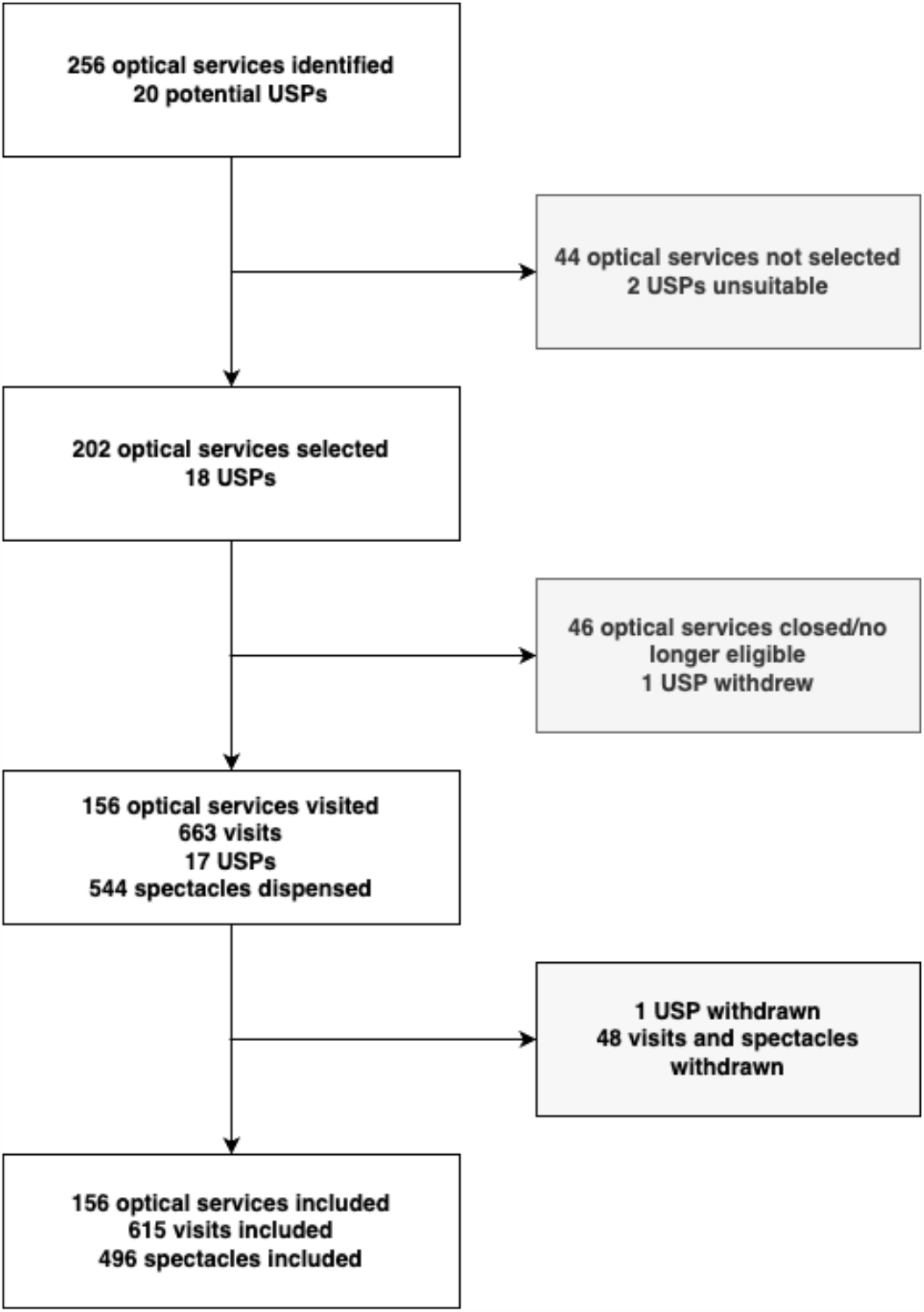
Flow chart of Unannounced Standardised Patient (USP) visits to optical services and dispensed spectacles

### Quality of spectacles and prescriptions

Of the 496 spectacles included, 259 (52.2%) were single-vision distance, and 237 (48.8%) were single-vision near, with 24 (4.8%) dispensed from public or non-governmental organization services and the remaining 472 from the private sector. Although a higher proportion of spectacles from public or non-governmental services were optimal (41.7%) versus private services (32.4%), this difference was non-significant (p=0.35). After weighting by population size, 35.1% (95%CI: 36.4-49.3%) of the spectacles were optimal quality, ranging from 20.3% in Siem Reap to 53.3% in Takeo (Table 1). The most common error observed in sub-optimal spectacles was horizontal prism outside of tolerance limits (96/333, 28.8%).

**Table 1:**
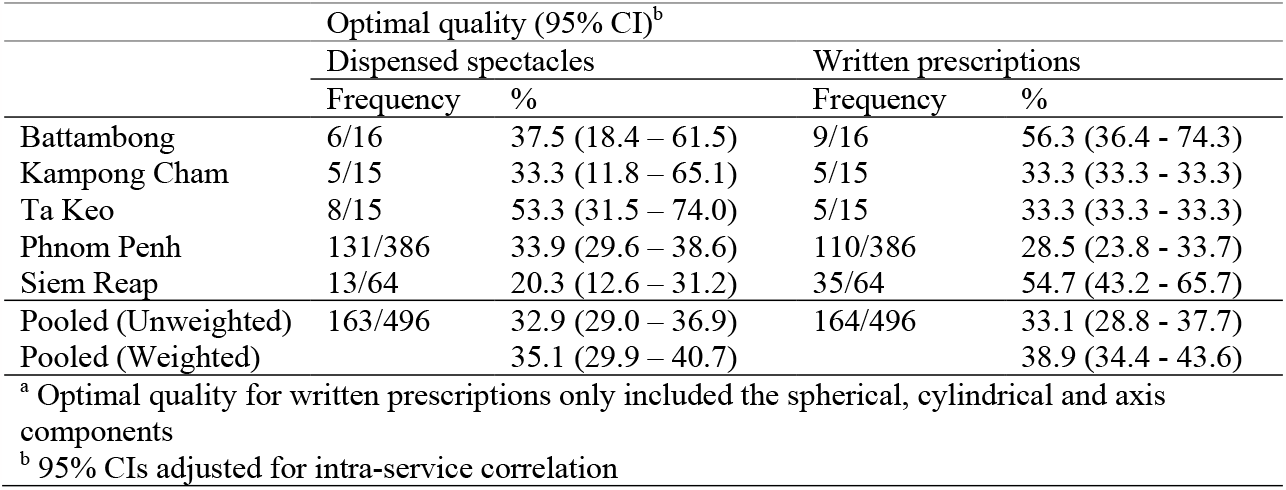
Percentage of dispensed spectacles and written prescriptions with optimal quality^a^

|  | Optimal quality (95% CI)<sup>b</sup> |  |  |  |
| --- | --- | --- | --- | --- |
|  | Dispensed spectacles |  | Written prescriptions |  |
|  | Frequency | % | Frequency | % |
| Battambang | 6/16 | 37.5 (18.4 – 61.5) | 9/16 | 56.3 (36.4 - 74.3) |
| Kampong Cham | 5/15 | 33.3 (11.8 – 65.1) | 5/15 | 33.3 (33.3 - 33.3) |
| Ta Keo | 8/15 | 53.3 (31.5 – 74.0) | 5/15 | 33.3 (33.3 - 33.3) |
| Phnom Penh | 131/386 | 33.9 (29.6 – 38.6) | 110/386 | 28.5 (23.8 - 33.7) |
| Siem Reap | 13/64 | 20.3 (12.6 – 31.2) | 35/64 | 54.7 (43.2 - 65.7) |
| Pooled (Unweighted) | 163/496 | 32.9 (29.0 – 36.9) | 164/496 | 33.1 (28.8 - 37.7) |
| Pooled (Weighted) |  | 35.1 (29.9 – 40.7) |  | 38.9 (34.4 - 43.6) |

<sup>a</sup> Optimal quality for written prescriptions only included the spherical, cylindrical and axis components
<sup>b</sup> 95% CIs adjusted for intra-service correlation

There was no significant difference in the proportion of distance and near spectacles that were of optimal quality (distance: 32.0% vs. near: 33.8%, p = 0.69). When comparing the individual spectacle components, a significantly lower proportion of distance spectacles were within tolerance limits for cylinder power (distance: 91.1% vs. near: 99.6%, p <0.001) and axis (distance: 43.1% vs. near: 100.0%, p <0.001) compared to near spectacles. However, no single-vision near spectacles were dispensed for those with astigmatism. A significantly lower proportion of near spectacles were within tolerance limits for horizontal prism compared to distance spectacles (distance: 90.3% vs. near: 48.1%, p <0.001).

On average, dispensed spectacles exhibited wider lens centres than the baseline pupillary distance for both distance (mean: 4.7mm, SD: 3.7mm) and near spectacles (mean: 6.8mm, SD: 4.4mm). The range fluctuated from 16.7mm wider to 2.3mm narrower than baseline for distance spectacles and 19.7mm wider to 0.3mm narrower for near spectacles.

Of the dispensed spectacles, 81.7% (n=405) matched the written prescription, and 38.9% (weighted, 95%CI: 34.4-43.6%) of the written prescriptions were within the tolerance limits for the sphere, cylinder, and axis components. Written prescriptions were inconsistent, particularly with the provision of pupillary distances. Over half of the written prescriptions (55.8%) did not provide any pupillary distance, and only 33 (6.7%) provided pupillary distance for both distance and near. For written prescriptions where only one pupillary distance was provided, sometimes it was unclear whether the value corresponded to distance or near.

### Optical service characteristics and associations with spectacle quality

Most clinicians providing refractive error care were men (82.9%). Autorefraction was the most common procedure (92.5%), while distance subjective refraction was absent in 35.9% of visits (Table 2).

**Table 2:** Distribution of testing procedures and associations with spectacle quality

| Testing procedure | Visits (N=615) | Dispensed spectacles (n=496) |  | p-value<sup>a</sup> |
| --- | --- | --- | --- | --- |
|  | Frequency (%) | Optimal quality frequency (%) | OR [95% CI] |  |
| Focimetry performed |  |  |  |  |
| No/Did not take spectacles | 342 (55.6) | 77/22733 (33.9) | 1.00 |  |
| Yes | 273 (44.4) | 86/269 (32.0) | 0.92 [0.65,1.30] | 0.618 |
| Distance VA checked at the beginning |  |  |  |  |
| No | 254 (41.3) | 65/206 (31.6) | 1.00 |  |
| Yes | 361 (58.7) | 98/290 (33.8) | 1.11 [0.76,1.60] | 0.590 |
| VA checked with pinhole |  |  |  |  |
| No | 537 (87.3) | 145/421 (34.4) | 1.00 |  |
| Yes | 78 (12.7) | 18/75 (24.0) | 0.60 [0.34,1.05] | 0.073 |
| Near VA checked at the beginning |  |  |  |  |
| No | 388 (63.1) | 95/297 (32.0) | 1.00 |  |
| Yes | 227 (36.9) | 68/199 (34.2) | 1.10 [0.77,1.58] | 0.591 |
| Autorefraction performed |  |  |  |  |
| No | 46 (7.5) | 13/38 (34.2) | 1.00 |  |
| Yes | 569(92.5) | 150/458 (32.8) | 0.94 [0.46,1.90] | 0.586 |
| Retinoscopy performed |  |  |  |  |
| No | 575 (93.5) | 155/458 (33.8) | 1.00 |  |
| Yes | 40 (6.5) | 8/38 (21.1) | 0.52 [0.25,1.10] | 0.087 |
| Distance subjective refraction |  |  |  |  |
| Spherical/cylinder power not checked | 318 (51.7) | 68/221 (30.8) | 1.00 |  |
| Only spherical component tested | 100 (16.3) | 43/89 (49.4) | 2.20 [1.31,3.68] | 0.003 |
| Only cylinder tested | 5 (0.8) | 0/5 (0.0) | NA |  |
| Sphere and cylinder tested | 192 (31.2) | 52/183 (28.4) | 0.89 [0.57,1.41] | 0.626 |
| Near VA with distance lenses checked |  |  |  |  |
| No | 432 (70.2) | 107/337 (31.8) | 1.00 |  |
| Yes | 183 (29.8) | 55/159 (35.2) | 1.17 [0.81,1.68] | 0.400 |
| Near subjective refraction performed |  |  |  |  |
| No | 345 (56.1) | 70/243 (28.8) | 1.00 |  |
| Yes | 270 (43.9) | 93/253 (36.8) | 1.44 [1.00,2.07] | 0.052 |
| Clinician used a phoropter |  |  |  |  |

|  | Visits (N=615) | Dispensed spectacles (n=496) |  |  |  |
| --- | --- | --- | --- | --- | --- |
| No | 257 (41.8) | 71/235<br>(30.2) | 1.00 |  |  |
| Yes | 137 (22.3) | 45/127<br>(35.4) | 1.27 | [0.81,1.97] | 0.293<sup>b</sup> |
| No distance subjective refraction | 221 (35.9) |  |  |  |  |
| Clinician used a trial frame |  |  |  |  |  |
| No | 29 (4.7) | 16/22<br>(72.7) | 1.00 |  |  |
| Yes | 365 (59.3) | 100/340<br>(29.4) | 0.16 | [0.07,0.36] | <0.001<sup>b</sup> |
| No distance subjective refraction | 221 (35.9) |  |  |  |  |
| Pupillary distance checked |  |  |  |  |  |
| No | 525 (85.4) | 129/410<br>(31.5) | 1.00 |  |  |
| Yes | 90 (14.6) | 34/86<br>(39.5) | 1.42 | [0.91,2.23] | 0.122 |

<sup>a</sup> Assessed via univariable logistic regression, accounting for within-optical service correlation
<sup>b</sup> Excludes 134 visits at which distance refraction was not performed, yet spectacles dispensed.

Surprisingly, the proportion receiving optimal spectacles was greater after only the spherical component of distance refraction had been performed compared to both spherical and cylindrical components. Conversely, more sub-optimal spectacles were dispensed following a trial frame refraction compared to not using a trial frame (Table 2). For most visits, communication was perceived as clear during eye examinations (84.7%), outcomes (91.5%), and spectacle recommendations (76.9%). Clear communication during eye examinations (OR [95%CI]: 5.00 [2.29-10.91], p<0.001) and outcomes (OR [95%CI]: 7.49 [1.63-34.31], p=0.01) was associated with receiving optimal spectacles.

After adjustment for USP refraction and stratification by spectacle type (distance vs near), receiving optimal distance spectacles was associated with having either myopia or hyperopia compared to having astigmatism alone or myopia astigmatism, and having focimetry performed and having only the cylinder component of distance subjective refraction testing (Table 3). Receiving optimal near spectacles was associated with having emmetropia with presbyopia compared to having hyperopia with presbyopia. No testing procedures were associated with receiving optimal near spectacles (Table 3).

**Table 3:**
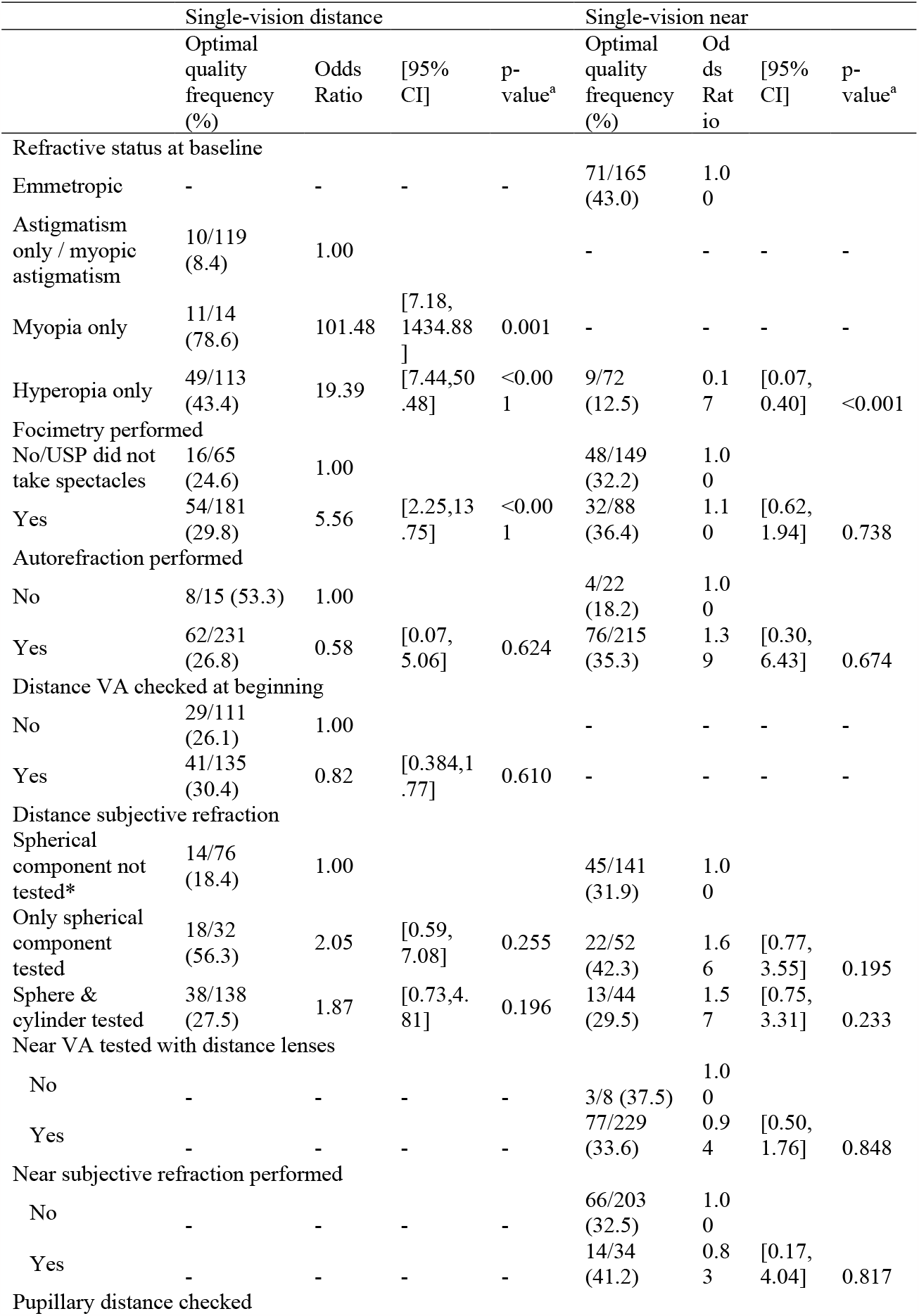
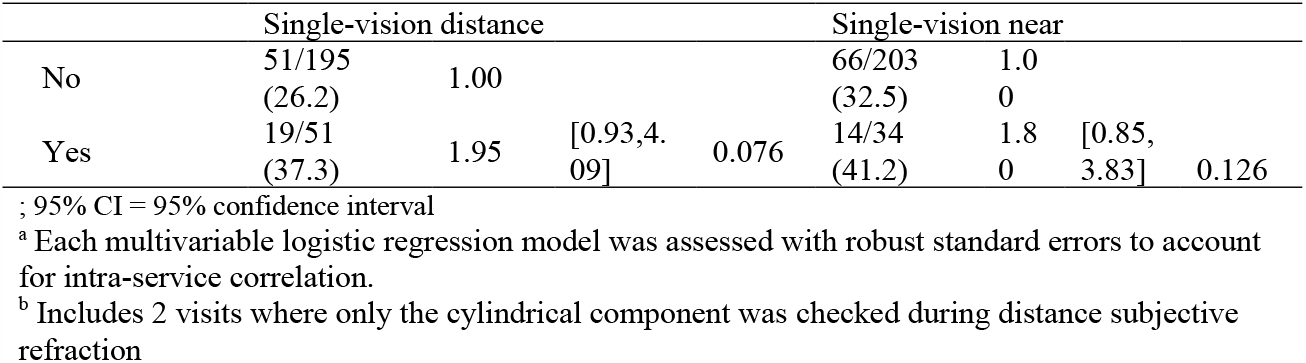
Association between testing procedures and spectacle quality among spectacle types (Single-vision distance, n=246; Single-vision near, n=237)

A total of 159 visits to 138 optical services were made by emmetropic individuals without presbyopia, with 44.0% (70/159) recommended spectacles and 46 dispensed (all in Phnom Penh). Men were less likely to receive unnecessary spectacle recommendations than women USPs (OR [95%CI]: 0.17 [0.06-0.489]; p<0.001). Spectacles were less likely to be recommended after checking distance (OR [95%CI]: 0.09 [0.04-0.20], p<0.001) and near VA (OR [95%CI]: 0.18 [0.06-0.49], p=0.001) at the beginning of the eye examination. Conversely, both spherical and cylindrical distance subjective refraction (OR [95%CI]: 2.90 [1.03-8.19]; p=0.04) and near subjective refraction (OR [95%CI]: 9.45 [4.23-21.14]; p<0.001) was associated with unnecessary spectacle recommendations.

### Vision and comfort

Good spectacle-corrected VA was achieved in 95.4% of distance spectacles (weighted percentage, 95% CI: 88.9-98.1%) for each eye separately, and 97.7% (weighted percentage, 95% CI: 94.4-99.1%) of near spectacles. More optimal distance spectacles achieved good distance VA compared to sub-optimal spectacles (100.0% vs 92.6%, p=0.01). A similar proportion achieved good near VA between optimal and sub-optimal single-vision near spectacles (99.3% vs 98.0%, p=0.44).

Discomfort or eyestrain was experienced in 47.3% (weighted, 95%CI: 38.8-56.0%) of distance spectacles, and at near in 35.1% of all spectacles (weighted, 95%CI: 31.1-39.3%). Fewer USPs experienced discomfort when wearing optimal spectacles compared to sub-optimal spectacles at distance (37.8% vs 55.7%, p=0.01) and near (16.8% vs 41.1%, p<0.001).

## Discussion

In this Q.REC Cambodia study, 35.1% of dispensed spectacles were optimal (weighted), 44.0% of emmetrope visits involved unnecessary prescription spectacle recommendations, and 18.3% of written prescriptions did not correspond with dispensed spectacles. Optimal spectacles were associated with effective communication and refractive status rather than testing procedures. Unnecessary spectacle recommendations were linked to increased subjective refraction testing and the absence of VA testing. Sex differences were observed, with men predominantly providing refractive error care and women more likely to be unnecessarily recommended spectacles.

Although an adequate level of VA was achieved with most sub-optimal spectacles, discomfort or eyestrain was much more prevalent with sub-optimal spectacles. This is likely to impact the frequency and duration of spectacle use in the community.

Inadequate dispensing seems to significantly contribute to sub-optimal spectacles, with horizontal prism being the most common issue. Only 14.6% of visits involved observed pupil distance measurement, and the average lens centres were wider than patients’ baseline pupil distance. This suggests that optical service staff may not be appropriately measuring pupil distance, or not accounting for convergence at near, which typically results in a narrower pupil distance. Additional training to enhance optical dispensers’ or mechanics’ skills to accurately measure pupil distances may reduce avoidable prismatic effects induced by spectacles.

The prevalence of astigmatism and hyperopia in Phnom Penh adults is estimated at 9.12% and 4.35% respectively.[4] Individuals with astigmatism or hyperopia combined with presbyopia were less likely to receive optimal spectacles. This highlights the need for improved refraction and/or dispensing techniques, particularly for those with complex prescriptions who are in greater need of appropriate spectacles.

In Cambodia, autorefraction is more commonly used than subjective refraction. Autorefraction is mainly a preliminary step in determining lens prescriptions, similar to subjective refraction. Studies have reported the inadequacy of autorefactors for prescribing glasses, [15 16] except for non-presbyopic adults in low-resource areas.[17 18] Retinoscopy is a more accurate alternative to autorefraction, but still requires complementary subjective methods for adult patients. [19]

Written prescriptions exhibited many inconsistencies, particularly with respect to pupillary distance. Standardised approaches to written spectacle prescriptions are needed and should include all parameters necessary for dispensers to make optimal spectacles.

Patient-centredness is vital for quality eye care. Ormsby et al. found that most adults were satisfied with their eye care when they received glasses at a public hospital.[20] Though our study didn’t measure satisfaction, it highlighted the importance of good communication for optimal spectacles. Communication skills include giving clear instructions, understanding patient needs, and informing them about their eye care. [21] Moreover, quality care involves avoiding unnecessary prescriptions. In our study, 44% of USPs who didn’t need glasses were prescribed them, with women being more likely to receive unnecessary prescriptions than men. This over-prescription could be due to insufficient refraction skills or an intent to increase sales –– the gender discrepancy warrants further investigation.

The recent Cambodian National Strategic Plan for Blindness Prevention & Control 2021 - 2030, emphasises strengthening integrated eye health service delivery.[22] The focus on enhancing primary eye care for those most in need, aligns with the pursuit of improved refractive error quality. The Cambodian Health Workforce Development Plan 2006-2015 acknowledges optometry and spectacle suppliers as integral to the healthcare system, especially in the private-for-profit sector. However, it notes a deficiency in regulation and registration in this sector, affecting service quality and workforce management. The plan mentions a seemingly balanced gender distribution in the health workforce, but lacks detailed breakdowns by sector or category. A new plan for 2021-2030 is in draft form.

This study has certain limitations that need to be acknowledged. Firstly, while visits were made to Kandal province, no spectacles were included in the primary analyses. Secondly, the Q.REC indicator does not currently have a minimum threshold to denote ‘high quality’, although it highlights various aspects of refractive error care that could improve. Thirdly, the Q.REC indicators are not adequate for comprehensive quality assessment of bifocal or progressive addition lenses. Finally, all near spectacles achieved good VA (with both eyes open and each eye separately), which may be attributable to the absence of small steps in the near VA charts when compared to logMAR distance charts.

Improving refractive error care in Cambodia is necessary, as only 35.1% of dispensed spectacles optimally address refractive error needs. Refraction and dispensing skills are currently limiting high-quality care in Cambodia, and there is a need for targeted clinical education, practice, and regulatory interventions. It is recommended that regulatory mandates are introduced to ensure optical services employ appropriately training and qualified staff. This measure would uphold professional standards and ultimately improve patient care. Additionally, there is a need to foster a diverse and inclusive workforce, with more opportunities for women to become integral members of the refractive error eye care. The evidence generated here can contribute to developing and implementing well-integrated eye health service delivery systems in line with the National Strategic Plan’s strategic priorities.

## Data Availability

Data are available upon reasonable request.

## Acknowledgements

We would like to thank Tokyo Bak for assistance in USP recruitment, training and data collection.

## Funding

This work was funded by The Fred Hollows Foundation with support from the Australia Government through the Australian NGO Cooperation Program (ANCP).

## Competing interests

LL and AB are consultants to The Fred Hollows Foundation.

## Patient consent for publication

Written informed consent was obtained from USPs. Optical services owners were informed of the study in writing and had the opportunity to opt-out verbally or in writing.

## Ethics approval

This research adhered to the tenets of the Declaration of Helsinki, and was approved by the University of New South Wales Human Research Ethics Committee (HC210102), and the National Ethics Committee for Health Research in Cambodia (043 NECHR).

## Data availability Statement

Data are available upon reasonable request.

## Notes

### Clinical Protocols

http://dx.doi.org/10.1136/bmjopen-2021-057594

