## Supplementary 1 for "Quality of Refractive Error Care in Cambodia: An Unannounced Standardised Patient Study"

Supplementary Material/Appendix 1: Criteria for optimally prescribed spectacles[10]

| Spectacle component | Tolerance limits* |
|---|---|
| Spherical power (in axis with most plus power) | ±0.50D |
| Cylindrical power | ±0.50D |
| Cylindrical axis (axis with most minus power)* | |
| If baseline \|cylinder power\| ≤ 0.50DC | ± 7 degrees |
| If baseline \|cylinder power\| > 0.50DC & ≤ 1.50DC | ± 5 degrees |
| If baseline \|cylinder power\| > 1.50DC | ± 2 degrees |
| Horizontal prism (total) | <1 prism dioptre |
| Vertical prism (total) | <0.5 prism dioptres |
